# Diabetes Diagnosis through Machine Learning: Investigating Algorithms and Data Augmentation for Class Imbalanced BRFSS Dataset

**DOI:** 10.1101/2023.10.18.23292250

**Authors:** Mohammad Mihrab Chowdhury, Ragib Shahariar Ayon, Md Sakhawat Hossain

**Affiliations:** Department of Mathematics and Statistics, Texas Tech University, Lubbock, TX, USA; Department of Electronics and Telecommunication Engineering, Rajshahi University of Engineering and Technology, Rajshahi, Bangladesh

**Keywords:** Diabetes, Imbalance Data, Machine Learning, Classification, Nominal Data

## Abstract

Diabetes is a prevalent chronic condition that poses significant challenges to early diagnosis and identifying at-risk individuals. Machine learning plays a crucial role in diabetes detection by leveraging its ability to process large volumes of data and identify complex patterns. However, imbalanced data, where the number of diabetic cases is substantially smaller than non-diabetic cases, complicates the identification of individuals with diabetes using machine learning algorithms. Our study focuses on predicting whether a person is at risk of diabetes, considering the individual’s health and socio-economic conditions while mitigating the challenges posed by imbalanced data. To minimize the impact of imbalance data, we employed several data augmentation techniques such as oversampling (SMOTE-N), undersampling (ENN), and hybrid sampling techniques (SMOTE-Tomek and SMOTE-ENN) on training data before applying machine learning algorithms. Our study sheds light on the significance of carefully utilizing data augmentation techniques, without any data leakage, in enhancing the effectiveness of machine learning algorithms. Moreover, it offers a complete machine learning structure for healthcare practitioners, from data obtaining to ML prediction, enabling them to make data-informed strategies.

## 1. Introduction

Communicable diseases like coronavirus, dengue fever, hepatitis, HIV/AIDS, and chickenpox have garnered global attention due to their potential for rapid cross-border transmission and profound impact on public health. Historically responsible for pandemics like Covid-19, Spanish flu, MARS, and cholera, these diseases have prompted extensive research and response efforts (Van Seventer and Hochberg, 2017; Kenworthy et al., 2018; Zumla et al., 2016; Green, 2015; Islam et al., 2021; Chowdhury et al., 2022). In contrast, non-communicable diseases (NCDs), including chronic illnesses, have received less recognition despite their significant global health implications (Budreviciute et al., 2020; Frumkin and Haines, 2019; WHO, Accessed March 22, 2023c). Rooted in genetic, physiological, environmental, and behavioral factors, NCDs progress gradually and are not contagious, often affecting specific populations within regions (Supakul et al., 2019; Bigna and Noubiap, 2019). Nonetheless, the prevalence of NCDs is surging, becoming the foremost cause of death and disability worldwide (Habib and Saha, 2010; CDC, Accessed March 24, 2023; WHO, Accessed March 22, 2023a; Divers et al., 2020). Diabetes, cardiovascular disease, chronic lung disease, and cancer are the predominant NCDs, collectively accounting for most NCD-related mortality (CDC, Accessed March 24, 2023; WHO, Accessed March 22, 2023b).

Diabetes, in particular, holds a prominent place within the NCD landscape. It ranks as the seventh leading cause of death in the United States (CDC, Accessed March 22, 2023e), with over 37.3 million Americans affected in 2019, approximately one in every ten people (CDC, Accessed March 22, 2023b,A). Alarmingly, many individuals with diabetes or prediabetes remain unaware of their condition, with every 1 in 5 people with diabetes and 8 in 10 people with pre-diabetes remaining undiagnosed (CDC, Accessed March 22, 2023b,A). Such underdiagnosis is concerning, given that individuals with diabetes are more vulnerable to seasonal and emerging diseases like COVID-19. Around 39.7% of hospitalized COVID-19 patients have diabetes as an underlying condition, rising to 46.5% for patients aged 50 to 64 (CDC, Accessed March 22, 2023d; Kastora et al., 2022; Rajpal et al., 2020).

Moreover, those with diabetes face a 60% higher risk of early mortality than those without diabetes and increased susceptibility to complications such as blindness, kidney failure, heart attacks, strokes, and limb amputation (CDC, Accessed March 24, 2023,A). Over the last two decades, diabetes prevalence has doubled in the USA, raising significant concerns (CDC, Accessed March 22, 2023c). Financially, diagnosed diabetes incurs substantial medical expenditures, averaging $16,752 per year, around $9,601 of which are attributed to diabetes, making it an economic burden. (Association, 2018; Chen et al., 2018; Association, Accessed June 22, 2023).

In this context, the timely identification of individuals at risk of diabetes is crucial for effective preventive measures. But mass testing for diabetes would be costly, time-consuming, and overwhelming for healthcare facilities. However, widespread diabetes testing entails significant costs, time consumption, and strains on healthcare resources. Machine learning emerges as a pivotal tool in diabetes detection, capitalizing on its capacity to process intricate datasets and discern complex patterns. Consequently, our study’s core objective is to comprehensively explore the intricate nexus between health, socioeconomic factors, and diabetes through accessible data and a machine-learning approach.

During the analysis phase, a notable challenge surfaced: a substantial class imbalance within the dataset, a significant hurdle for achieving accurate results via machine learning algorithms. Existing literature underscores the adverse impact of dataset imbalance on algorithm performance, with established methods demonstrating subpar performance in identifying minority classes when data leakage is absent (Fernández et al., 2018; Kaur et al., 2019; Ul Hassan et al., 2022). We employed various sampling techniques and ensemble machine learning algorithms to address this imbalance (Chawla et al., 2002; Anand et al., 2010), ensuring no data leakage occurred at any study stage. Valuing these techniques and identifying the most effective strategy for handling imbalanced data within the context of machine learning algorithms formed critical dimensions of our investigation. Impressively, all sampling techniques yielded superior recall values compared to the original dataset. Our methodology notably highlights the superior performance of the Editest Nearest Neighbors (ENN) sampling technique across all metrics. Moreover, our findings align with existing research, indicating that higher BMI, elevated blood pressure levels, and advanced age correlate with elevated diabetes risk (Leong and Wilding, 1999; Gray et al., 2015; Group, 2010; Geiss et al., 2002; Caspersen et al., 2012; Ahima, 2009; Morley, 2008).

## 2. Study Design

The step-by-step workflow, as illustrated in Diagram Figure 1, outlines the various stages and processes involved in conducting our study. The diagram provides a visual representation of how the study progresses from data collection to analysis and interpretation of results.

**Figure 1:**
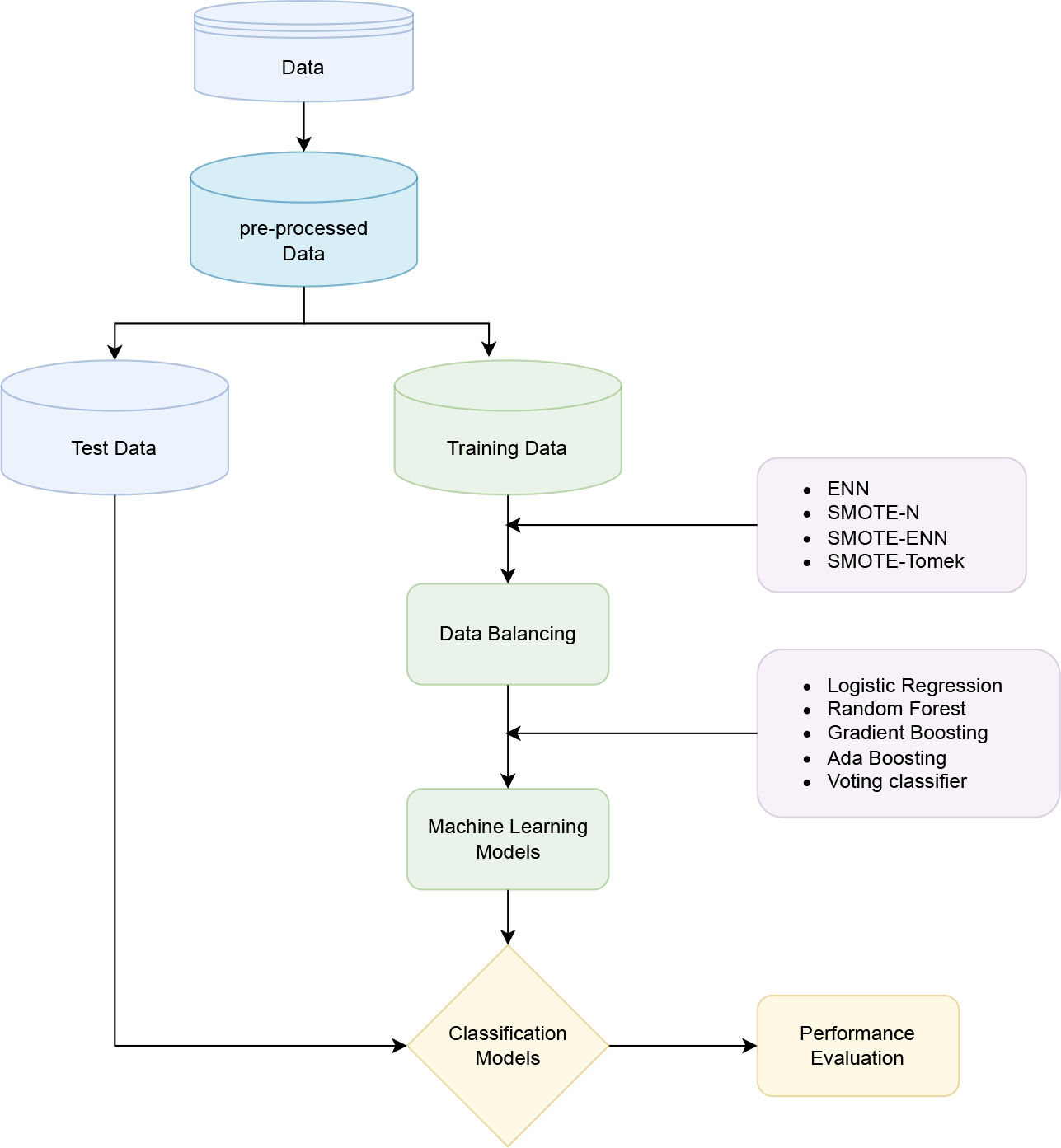
Illustration of the Study Process from Data Collection to Result Interpretation.

### 2.1. Data Overview

This study employs the 2021 - Behavioral Risk Factor Surveillance System (BRFSS) dataset, sourced from telephone surveys, encompassing USA residents’ health behaviors, conditions, and socioeconomic aspects (CDC, Accessed March 22, 2023a). The BRFSS-2021 dataset holds 438,693 records featuring 303 attributes. For diabetes classification, our binary approach excludes gestational diabetes, focusing on Type 1 and Type 2 diabetes (CDC, Accessed March 22, 2023g,A; Association, 2010). The latter is omitted due to its transient nature, which is linked explicitly to pregnancy (Buchanan et al., 2005). Gestational diabetes is reversible; it typically resolves after childbirth, unlike Type 1 and Type 2 diabetes, which require lifelong management. Type 1 diabetes results from immune-driven insulin deficiency (Katsarou et al., 2017; Eisenbarth, 1986), while Type 2 diabetes emerges from insulin resistance (Astrup and Finer, 2000; Chatterjee et al., 2017). Approximately 90-95% of cases are Type 2, while 5-10% are Type 1 (Association, 2010; CDC, Accessed March 22, 2023g,A). This study adopts a simplified binary categorization approach by grouping Type 1 and Type 2 diabetes under the variable “diabetes”.

#### 2.1.1. Data Preprocessing

We meticulously curated the most pertinent features for our study through an exhaustive review of existing literature (Robertson et al., 2011; Dinh et al., 2019; Hill-Briggs et al., 2021; Shriraam et al., 2021; Asiimwe et al., 2020; Budreviciute et al., 2020; Supakul et al., 2019; Ullah et al., 2022), resulting in a selection of 20 variables. Our independent variables encompass BMI, AGE, Income, Smoking, Blood Pressure, Cholesterol, Heart Disease, Asthma, Kidney Disease, Marital Status, Education, General Health Condition, Exercise, Arthritis, Depression, Food and Vegetable Consumption, Sex, and Diabetes as the dependent variable. The target variable inquired whether respondents had ever been told they had diabetes, with response options including Yes (1), Yes (But female told only during pregnancy) (2), No (3), No (Pre-diabetes or borderline diabetes) (4), Don’t know/Not Sure (7), Refused (9), and Blank (Table 1).

**Table 1:** Diabetes Classification Question and Response.

| Target Question | Response | ID |
| --- | --- | --- |
| "Ever told you had diabetes?" | Answered "yes" | 1 |
|  | Answered "yes" but, "Female told only during pregnancy" | 2 |
|  | No (Diabetes) | 3 |
|  | No (Pre-diabetes or borderline diabetes) | 4 |
|  | Don't know/Not Sure | 7 |
|  | Refused | 9 |

To align the dataset for machine learning analysis, we conducted preprocessing by eliminating missing values and disregarding instances where respondents indicated “Don’t Know/Not Sure,” “Refused,” or left fields “Blank.” Furthermore, we excluded gestational diabetes due to its temporary and reversible nature. Pregnant women were also removed from our study set to mitigate biases. Consequently, our study considered only yes (1) without gestational diabetes and no (3) without pregnant women. The dataset’s diabetic and non-diabetic percentages are presented in Table 2, with age-specific distributions in Table 3. Notably, the dataset demonstrates a minimal impact of diabetes on young individuals, aligning with broader literature indicating increased diabetes risk for individuals over 45 years of age (CDC, Accessed March 22, 2023e). Consequently, we focused on individuals aged 40 and above to harmonize our dataset with existing literature and address dataset imbalances. This dataset preparation culminated in a dataset size of (262,958, 21), providing a solid foundation for subsequent data preprocessing and analysis.

**Table 2:** Structure of the Dataset.

| Type | Size | Ratio |
| --- | --- | --- |
| Diabetic | 48,581 | 15% |
| Non-Diabetic | 265,332 | 85% |
| Total | 313,913 | 100% |

**Table 3:** Age Structure of The Dataset.

| Age | Criteria | Percentage | Age | Criteria | Percentage |
| --- | --- | --- | --- | --- | --- |
| 18 - 24 | Diabetic<br>Non-Diabetic | 0.02<br>0.98 | 55 - 59 | Diabetic<br>Non-Diabetic | 0.17<br>0.83 |
| 25 - 29 | Diabetic<br>Non-Diabetic | 0.02<br>0.98 | 60 - 64 | Diabetic<br>Non-Diabetic | 0.19<br>0.81 |
| 30 - 34 | Diabetic<br>Non-Diabetic | 0.03<br>0.97 | 65 - 69 | Diabetic<br>Non-Diabetic | 0.20<br>0.80 |
| 35 - 39 | Diabetic<br>Non-Diabetic | 0.05<br>0.95 | 70 - 74 | Diabetic<br>Non-Diabetic | 0.23<br>0.77 |
| 40 - 44 | Diabetic<br>Non-Diabetic | 0.07<br>0.93 | 75 - 79 | Diabetic<br>Non-Diabetic | 0.24<br>0.76 |
| 45 - 49 | Diabetic<br>Non-Diabetic | 0.11<br>0.89 | 80 or older | Diabetic<br>Non-Diabetic | 0.20<br>0.80 |
| 50 - 54 | Diabetic<br>Non-Diabetic | 0.13<br>0.87 | Don't know/<br>Refused/ Missing | Diabetic<br>Non-Diabetic | 0.13<br>0.87 |

#### 2.1.2. Splitting the dataset

The organized dataset underwent partitioning into training and testing subsets, utilizing an 80% training and 20% testing ratio. For this purpose, we employed Python’s model selection library’s default test-train split command. This command not only shuffles the dataset but also employs a stratified approach, thereby preserving the proportional representation of each Diabetes class observed in the original dataset (See in Figure 2 and Table 4).

**Table 4:** Before and After Splitting.

| Type | Size | Percentage | Catagory | Type | Size | Percentage |
| --- | --- | --- | --- | --- | --- | --- |
| Diabetic<br>Non-Diabetic | 46,944<br>216,014 | 18%<br>82% | Train | Diabetic<br>Non-Diabetic | 37,456<br>172,910 | 18%<br>82% |
| Total | 262,958 | 100% | Test | Diabetic<br>Non-Diabetic | 9,488<br>43,104 | 18%<br>82% |

**Figure 2:**
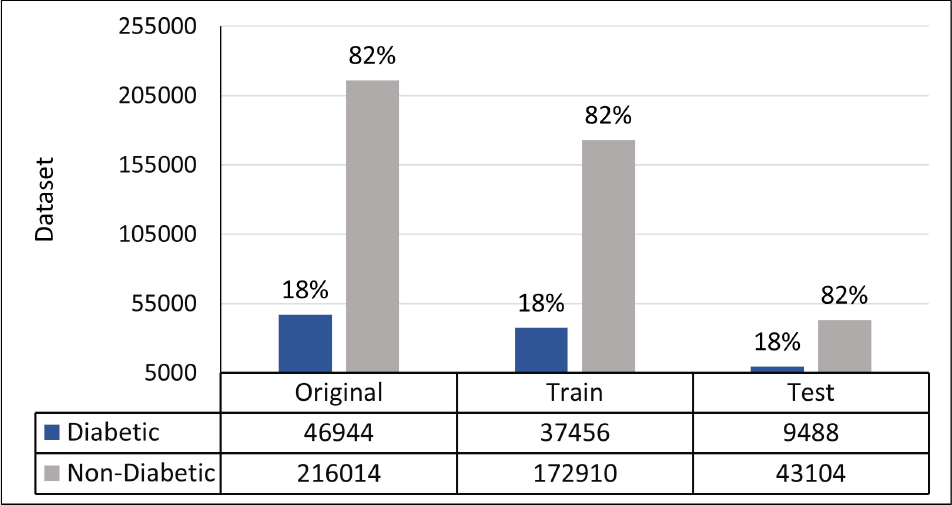
Visual Representation of Before and After Splitting the Dataset

### 2.2. Balancing Techniques

In Table 4, the imbalanced nature of our dataset’s target variable is evident, with diabetic patients representing only 18.0%. To address this class imbalance, we implemented four distinct sampling techniques on the training data tailored to nominal data. These techniques encompass oversampling, undersampling, and hybrid approaches. We applied these techniques to balance the dataset and enhance our models’ performance.

For the oversampling technique, we adopted the Synthetic Minority Over-sampling Technique for nominal data (SMOTE-N) (Chawla et al., 2002), given the nominal nature of our data. As for undersampling, we utilized the Edited Nearest Neighbours algorithm (Alejo et al., 2010). We employed SMOTE-Tomek, which combines SMOTE-N with Tomek Links, and SMOTE-ENN, which integrates SMOTE-N with Edited Nearest Neighbours (Chawla et al., 2002) as hybrid techniques. The specific details of these sampling techniques are briefly outlined in the following sections.

#### 2.2.1. SMOTE (Synthetic Minority Over-sampling Technique)

SMOTE is a widely acknowledged technique for addressing imbalanced data in classification problems (Chawla et al., 2002; Burez and Van den Poel, 2009; He and Garcia, 2009). It operates by generating synthetic instances for the minority class through interpolation between existing examples rather than through replacement-based oversampling (Chawla et al., 2002). For each minority instance *x*_*i*_, SMOTE constructs N synthetic examples by interpolating with its K nearest neighbors *x*_*j*_, incorporating a parameter *λ* within the range of 0 to 1. This interpolation process is represented as *x*_*new*_ = *x*_*i*_ + *λ*(*x*_*j*_ *− x*_*i*_), where *x*_*i*_ and *x*_*j*_ denote feature value vectors.

#### 2.2.2. ENN (Edited Nearest Neighbour)

ENN, or Edited Nearest Neighbors, is an enhanced classification algorithm derived from k-Nearest Neighbors (k-NN), designed to eliminate noisy and mislabeled instances from the training dataset. This refinement improves classification accuracy, rendering ENN particularly valuable for addressing imbalanced datasets (Alejo et al., 2010; Wilson, 1972).

#### 2.2.3. SMOTE-ENN and SMOTE-TOMEK

SMOTE-ENN emerges as the fusion of SMOTE and ENN techniques, effectively tackling imbalanced data classification challenges. By merging SMOTE’s minority class over-sampling with ENN’s majority class undersampling, the method harmonizes the dataset’s distribution. This technique was formulated by (Batista et al., 2004) as a robust approach for handling class imbalance.

Similarly, SMOTE-TOMEK is another amalgamation technique employed in imbalanced data classification scenarios. It is a potent strategy renowned for its efficacy in addressing imbalanced datasets, particularly when faced with noisy or overlapping instances. By integrating SMOTE and Tomek Links, this technique effectively navigates imbalanced scenarios to improve model performance.

### 2.3. Encoding nominal values

Given that all dataset features are of nominal type, using the values directly would mislead machine learning models. Due to this, we applied to the dataset one-hot encoding. This approach created distinct categories based on unique values in each column, subsequently expanding the number of columns to 92.

### 2.4. Machine Learning Algorithms

Employing a diverse set of machine learning algorithms, including Logistic Regression, Gradient Boosting, AdaBoost, and Random Forest classifiers, we strived to leverage their distinct strengths for enhanced predictions. The top three performing algorithms were aggregated using a voting classifier to optimize overall predictive accuracy for each sampling technique. This collaborative approach aimed to bolster the accuracy and robustness of our predictions across different sampling strategies. The following section provides a concise overview of these algorithmic components.

#### 2.4.1. Logistic Regression

Logistic Regression (LR) models binary responses using a set of independent predictors. Let *P*_*i*_ denote the probability of a patient responding “Yes” and 1 *− P*_*i*_ the probability of responding “No”. The LR model equation is formulated as follows:

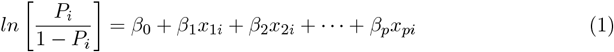

This equation computes the natural logarithm of the odds ratio of responses, with *β*_0_, *β*_1_, *β*_2_, *…, β*_*p*_ representing the model coefficients. Subsequently, the equation is transformed as:

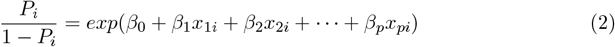

This further leads to:

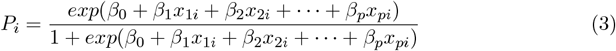

#### 2.4.2. Random Forest

Initially introduced by (Breiman, 2001), Random Forest operates as a tree-based ensemble prediction model. This prediction model builds multiple decision trees using randomly selected predictor variables and training datasets. The independent variables are represented as *X* = (*X*_1_, *X*_2_, …, *X*_*k*_), while *Y* signifies the response variable. The main goal is to predict *Y* by establishing a prediction function *f* (*X*) ((Cutler et al., 2012)).

The model minimizes the loss function *L*(*Y, f* (*X*)) to determine the prediction function. In classification tasks, the commonly used loss function is the zero-one loss.

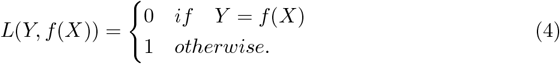

#### 2.4.3. Gradient Boosting

Gradient Boosting, a widely adopted ensemble method for classification and regression tasks, was initially introduced by (Friedman, 2001). This technique constructs an additive model by sequentially incorporating weak learners, enhancing the model’s overall performance ((Hastie et al., 2009)). By iteratively focusing on the mistakes made by prior learners, each subsequent learner aims to correct and improve upon the prior ones, effectively creating a powerful ensemble model that effectively leverages each learner’s strengths.

#### 2.4.4. AdaBoost (Adaptive Boosting)

AdaBoost, introduced by (Freund and Schapire, 1997), is an iterative algorithm designed to enhance the performance of weak classifiers, also known as base classifiers. AdaBoost improves data classification capabilities by adapting these base classifiers’ errors iteratively. This process contributes to a reduction in both bias and variance. The algorithm’s strength lies in its ability to continuously train and refine the classifiers, resulting in an ensemble model that leverages the individual strengths of these classifiers, ultimately yielding improved overall classification accuracy (Wu et al., 2020).

#### 2.4.5. Voting Classifier

Voting classifiers belong to the ensemble machine learning category, where the outcomes of multiple distinct classifiers are combined to yield a final prediction (Beyeler, 2017). This approach harnesses collective knowledge to bolster prediction accuracy and reliability. By amalgamating predictions from each classifier, the voting classifier employs a majority rule mechanism to make a final prediction, opting for the class label that accumulates the highest number of votes. This collaborative approach enhances the model’s overall predictive power, benefiting from its constituent classifiers’ diverse insights.

### 2.5. Evaluation Metrics

During our evaluation, we focused on four essential metrics: precision, recall, accuracy, and AUC-ROC (Japkowicz, 2006). In the context of imbalanced data, recall gains significance by accurately pinpointing positive instances within the minority class, which is crucial in real-world applications. Unlike accuracy, which can be deceptive in such datasets, robust recall guarantees adept detection and prediction of minority class instances. This section briefly elaborates on the evaluation metrics’ descriptions.

#### 2.5.1. Precision

Precision addresses how accurate identifications are, expressed as the percentage of correct optimistic predictions out of all predicted positives. Also known as positive predictive value (PPV), it is calculated by dividing true positives by the sum of true positives and false positives.

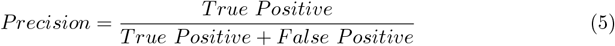

#### 2.5.2. Recall (Sensitivity)

Recall quantifies the proportion of true positives correctly detected, reflecting the ratio of true positives to all instances that should have been identified as positive. In binary classification, recall corresponds to sensitivity.

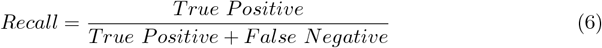

#### 2.5.3. Accuracy

Accuracy gauges the percentage of correct classifications achieved by a trained model, calculated by dividing the sum of true negatives and true positives by the total instances in the dataset. This metric is particularly effective for balanced classification tasks with relatively even class representation.

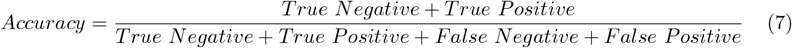

#### 2.5.4. AUC-ROC

The Area Under the Receiver Operating Characteristic Curve (AUC-ROC) evaluates binary classification model performance. It measures the model’s ability to differentiate between positive and negative classes by plotting True Positive Rate (TPR) against False Positive Rate (FPR). A perfect classifier yields an AUC of 1.0, while a random classifier has an AUC of 0.5. The AUC-ROC value is computed by integrating the TPR-FPR curve over the entire FPR range from 0 to 1.

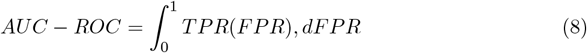

## 3. Hyper Parameter Tuning

To optimize model performance, we leveraged GridSearchCV for hyperparameter tuning on the training data. This method systematically explores various hyperparameter combinations within a predefined grid, enabling the models to be trained and assessed for the most favorable configuration that maximizes performance metrics. Through this approach, we fine-tuned our models, identifying optimal hyperparameters that enhanced accuracy and better generalization when handling unseen data.

## 4. Result

Diabetes, a chronic condition affecting millions globally, arises from the body’s inability to regulate blood sugar levels, resulting in elevated glucose levels in the bloodstream and severe complications, including nerve and kidney damage, vision issues, and cardiovascular disease. Timely identification of individuals at risk of developing diabetes is essential for effective intervention. Traditional testing methods for diabetes, while valuable, can be costly, time-consuming, and may not capture the full complexity of risk factors. In contrast, machine learning offers distinct advantages by leveraging existing data to provide costeffective and efficient solutions, particularly crucial for early detection, where subtle risk factors might play a significant role. However, the challenge of imbalanced data, where the number of diabetes cases is limited compared to non-diabetic instances, poses difficulties in training machine learning algorithms. Overcoming this hurdle is essential for accurate diabetes prediction using machine learning algorithms.

Our study focuses on comparing three sampling techniques: oversampling, undersampling, and hybrid methods (Figure 3,Figure 4) to address data imbalance. We apply the Synthetic Minority Over-Sampling Technique (SMOTE-N) to oversample the minority class (individuals with diabetes), creating a more balanced dataset for machine learning algorithms to learn. For undersampling, we utilize Editing Nearest Neighbour to reduce the number of samples in the majority class (people without diabetes) and mitigate the dataset’s imbalance. Furthermore, we explore hybrid techniques, including SMOTE-Tomek and SMOTE-ENN, to achieve a balanced dataset.

**Figure 3:**
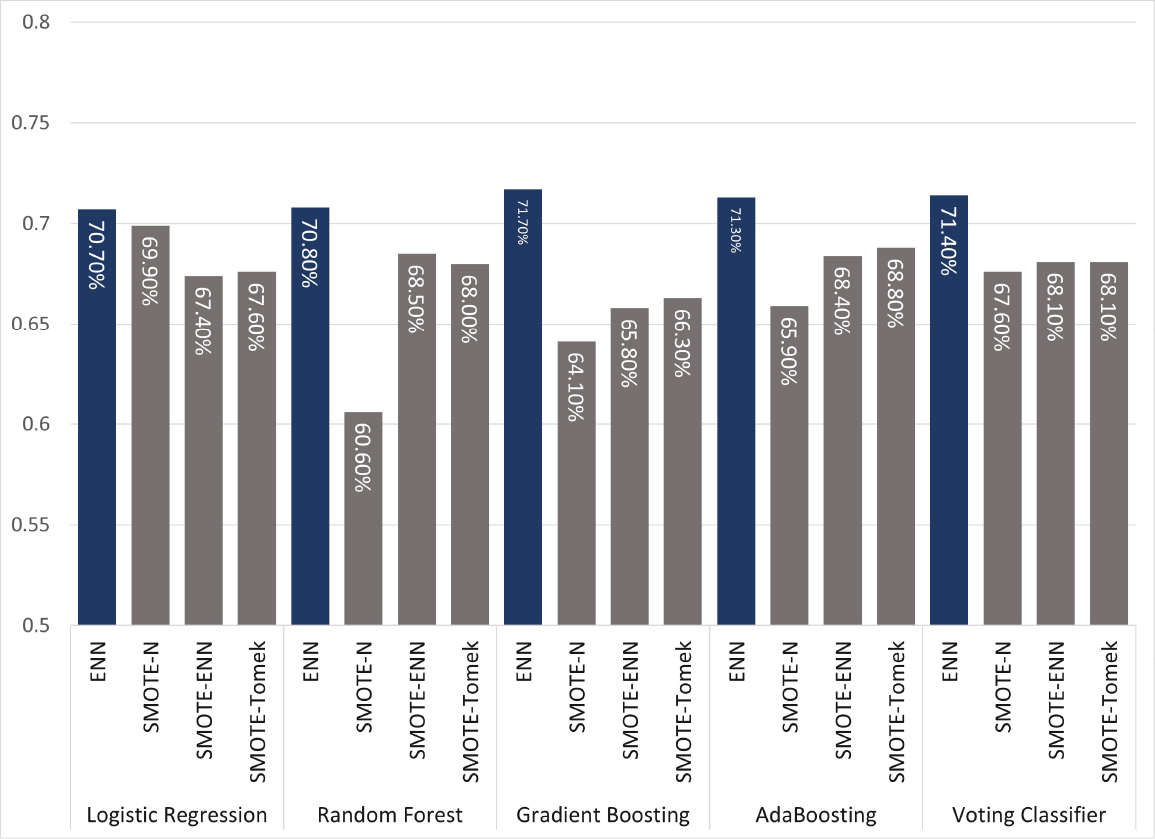
Recall for Different Sampling Techniques: Edited Nearest Neighbors (ENN), Synthetic Minority Over-sampling Technique (SMOTE-N), SMOTE-Tomek, and SMOTE-ENN for Logistic Regression, Random Forest, Gradient Boosting, and AdaBoost

**Figure 4:**
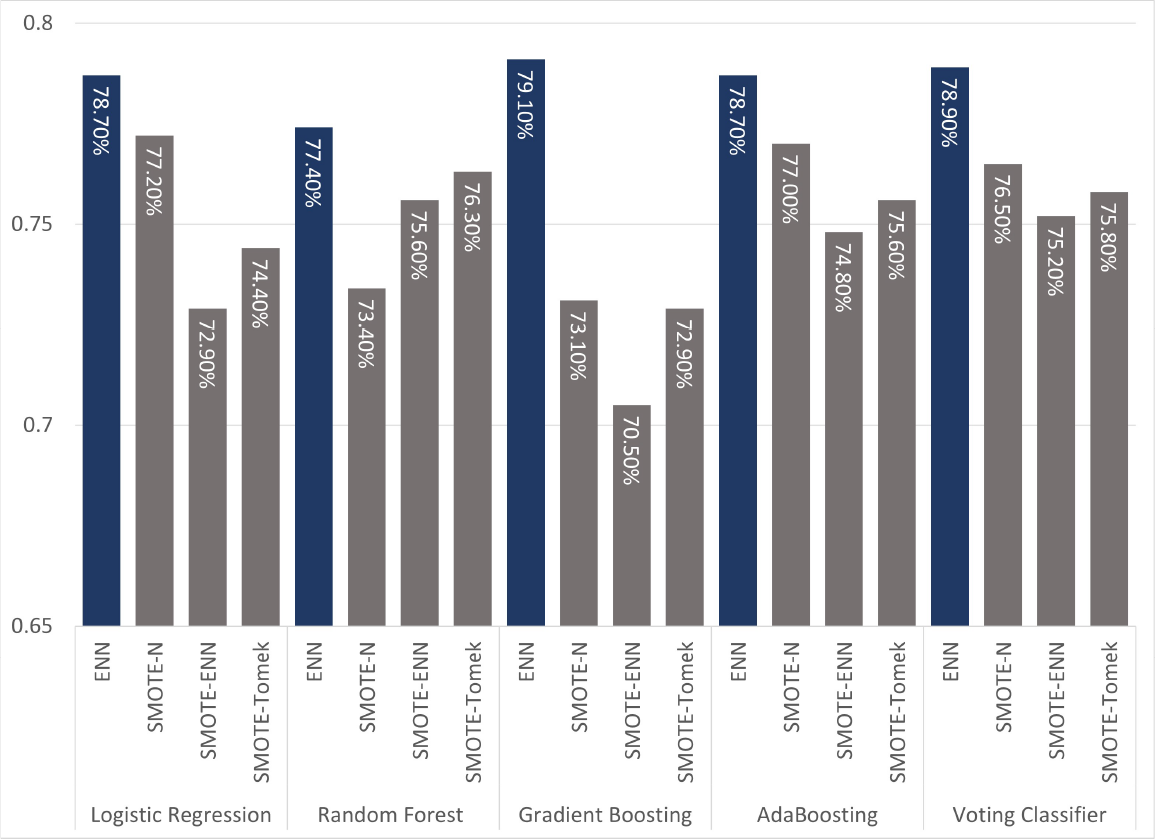
AUC for Different Sampling Techniques: Edited Nearest Neighbors (ENN), Synthetic Minority Over-sampling Technique (SMOTE-N), SMOTE-Tomek, and SMOTE-ENN for Logistic Regression, Random Forest, Gradient Boosting, and AdaBoost

Our study employs several machine learning algorithms on each sampling technique, including Logistic Regression, Random Forest, AdaBoost, and Gradient Boost (Table 6). Following an initial evaluation of AUC scores and recall metrics, we fine-tune hyperparameters using GridSearchCV for these algorithms, utilizing the TTU High-Performance Computing Center (HPCC) for computational efficiency. Model performance evaluation includes precision, recall, accuracy, and AUC scores (Table 6,Figure 4). Specifically, our focus lies on recall, as it is paramount in disease detection due to the inherent imbalance of positive cases(Figure 3) (Japkowicz, 2006).

The challenge of imbalanced data becomes evident when comparing the model performance before and after applying the sampling techniques (Figure 5). Applying Logistic Regression, Random Forest, Gradient Boosting, and AdaBoosting on the raw data reveals relatively high accuracy scores, ranging from 81.7% to 83.0%. However, this high accuracy is accompanied by relatively low recall scores, ranging from 57.4% to 58% (Table 5). This discrepancy underscores the misclassification of positive cases as negative, adversely affecting timely diagnosis and patient care. Misclassification can lead to missed interventions and an increased risk of complications.

**Table 5:** Before implementing the data balancing techniques.

| Models | Precision | Recall | Accuracy | AUC |
| --- | --- | --- | --- | --- |
| Logistic Regression | 0.718 | 0.577 | 0.83 | 0.787 |
| Random Forest | 0.663 | 0.574 | 0.817 | 0.754 |
| Gradient Boosting | 0.722 | 0.575 | 0.83 | 0.789 |
| AdaBoosting | 0.717 | 0.58 | 0.83 | 0.787 |

**Figure 5:**
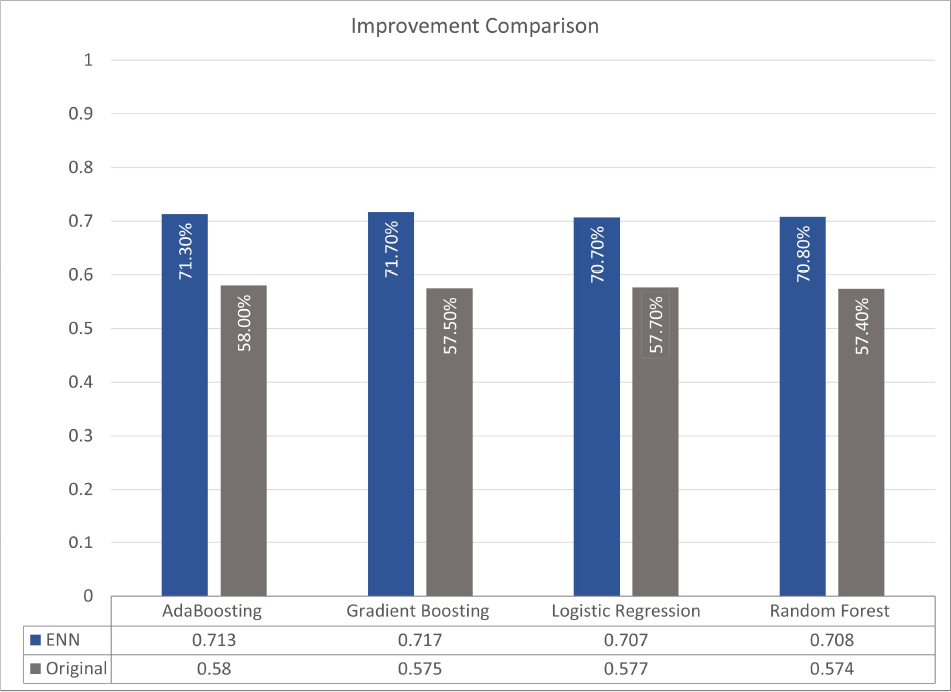
Comparison of improvement of recall value between original and best sampling technique (ENN) for different ML algorithms

**Figure 6:**
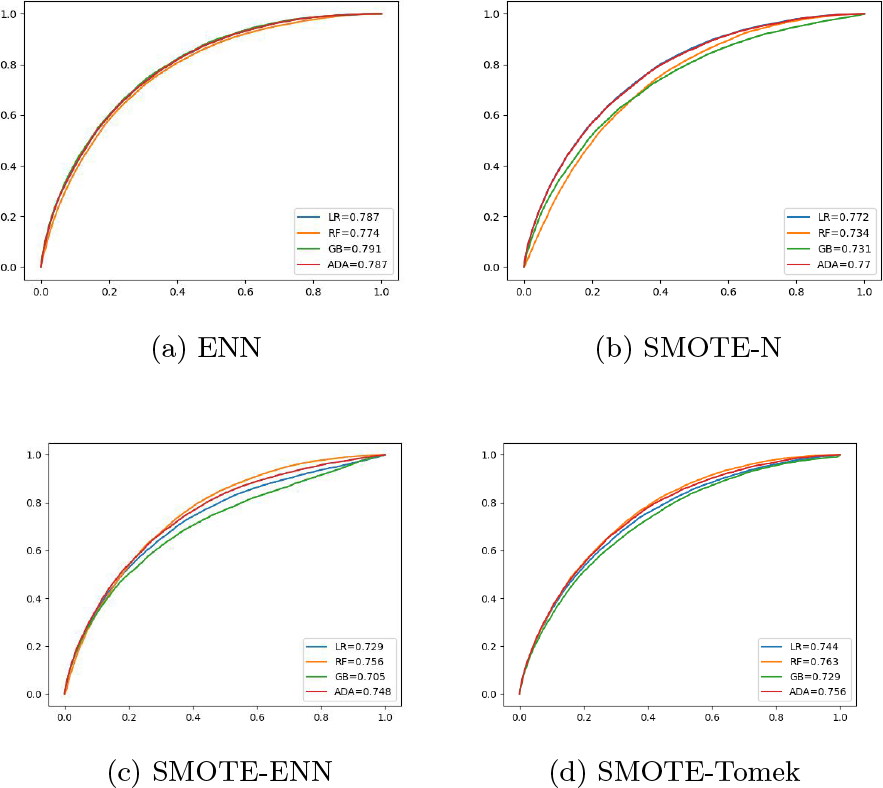
Comparative Evaluation of Area Under the Receiver Operating Characteristic (AUC-ROC) Curves for Different Sampling Techniques: Edited Nearest Neighbors (ENN), Synthetic Minority Over-sampling Technique (SMOTE-N), SMOTE-Tomek, and SMOTE-ENN for Logistic Regression, Random Forest, Gradient Boosting, and AdaBoost

Our study leverages various data-balancing techniques to address these limitations, resulting in substantial improvements, particularly in recall (Table 6). Among the strategies, the ENN undersampling technique stands out, enhancing recall by approximately 14.2% when paired with Gradient Boosting. AdaBoosting and Logistic Regression, paired with ENN, also yield significant recall improvements of 13.3% and 13.1%, respectively (Figure 5). The effectiveness of these techniques emphasizes the importance of data balancing, particularly for minority class identification.

**Table 6:** After implementing different sampling strategies.

| Sampling Strategy | Models | Precision | Recall | Accuracy | AUC |
| --- | --- | --- | --- | --- | --- |
| ENN | Logistic Regression | 0.623 | 0.707 | 0.625 | 0.787 |
|  | Random Forest | 0.629 | 0.708 | 0.691 | 0.774 |
|  | Gradient Boosting | 0.635 | 0.717 | 0.703 | 0.791 |
|  | AdaBoosting | 0.632 | 0.713 | 0.694 | 0.787 |
| SMOTE-N | Logistic Regression | 0.627 | 0.699 | 0.706 | 0.772 |
|  | Random Forest | 0.618 | 0.606 | 0.783 | 0.734 |
|  | Gradient Boosting | 0.63 | 0.641 | 0.775 | 0.731 |
|  | AdaBoosting | 0.652 | 0.659 | 0.791 | 0.770 |
| SMOTE-ENN | Logistic Regression | 0.609 | 0.674 | 0.679 | 0.729 |
|  | Random Forest | 0.624 | .685 | 0.718 | 0.756 |
|  | Gradient Boosting | 0.602 | 0.658 | 0.684 | 0.705 |
|  | AdaBoosting | 0.61 | 0.684 | 0.642 | 0.748 |
| SMOTE-Tomek | Logistic Regression | 0.617 | 0.676 | 0.710 | 0.744 |
|  | Random Forest | 0.63 | 0.68 | 0.742 | 0.763 |
|  | Gradient Boosting | 0.613 | 0.663 | 0.719 | 0.729 |
|  | AdaBoosting | 0.623 | 0.688 | 0.711 | 0.756 |

Our study extends to using ensemble methods, such as the soft voting classifier, to achieve a robust and stable model by combining top-performing algorithms based on recall. For instance, when employing the ENN sampling method, the voting classifier attains a recall of 71.4% and an AUC of 78.9% (Table 7), highlighting its capability to identify positive cases and differentiate between classes more precisely. Other techniques, including SMOTE-ENN and SMOTE-N, yield promising results, further underscoring the value of ensemble methods for enhancing model accuracy in imbalanced datasets.

**Table 7:** Voting Classifier.

| Sampling Strategy | Precision | Recall | Accuracy | AUC |
| --- | --- | --- | --- | --- |
| ENN | 0.635 | 0.714 | 0.706 | 0.789 |
| SMOTE-N | 0.635 | 0.676 | 0.757 | 0.765 |
| SMOTE-ENN | 0.617 | 0.681 | 0.699 | 0.752 |
| SMOTE-Tomek | 0.625 | 0.681 | 0.728 | 0.758 |

Our study identifies essential risk factors for diabetes classification through feature selection and analysis. Key factors include age, BMI, and blood pressure, consistent with prior research. We emphasize that machine learning techniques offer a significant advantage over traditional testing methods because they leverage existing data for cost-effective and efficient early detection. Our findings indicate that while under-sampling techniques exhibit superior outcomes, hybrid techniques provide comparable results, indicating their potential for effective diabetes classification. Computational resources may be limited. Even running logistic regression can provide valuable initial insights. Ultimately, our study underscores the pivotal role of machine learning in enhancing early diabetes detection using existing data, given the challenges posed by imbalanced datasets. Further hyperparameter tuning for computationally intensive algorithms and larger datasets may yield optimized results. Ultimately, our study underscores the pivotal role of machine learning in enhancing early diabetes detection using existing data, given the challenges posed by imbalanced datasets.

## 5. Conclusion

The implications of our study carry profound significance within the context of the prevailing global burden of diabetes. With millions of individuals affected worldwide, diabetes has substantial social, economic, and healthcare ramifications. According to data from the International Diabetes Federation, an estimated 463 million adults between the ages of 20 and 79 were grappling with diabetes in 2019, and this number is projected to escalate to an astounding 700 million by 2045 (Saeedi et al., 2019). Beyond the personal toll of diabetes on physical health and well-being, the disease significantly strains healthcare systems and economies globally. It is predicted that annual healthcare expenditures linked to diabetes will surpass $800 billion by 2045 (Williams et al., 2020).

In light of this pressing issue, the imperative of proactive diabetes prevention and effective management strategies becomes clear. This necessitates the identification and mitigation of pivotal risk factors for the disease and the refinement of diagnostic and treatment approaches. In this context, machine learning algorithms and data augmentation techniques play a critical role in enhancing accurate and efficient predictions, shedding light on the intricate web of underlying causes and enabling early identification of individuals at risk of diabetes. Our study delves into the application of data augmentation techniques such as SMOTE-N, SMOTE-ENN, SMOTE-Tomek, and ENN, with a specific focus on the training dataset. Our findings highlight the potential effectiveness of these techniques, as they yield more precise and potent machine-learning models when employed wisely. However, it is imperative to be aware of the potential pitfalls associated with these techniques. For instance, indiscriminate application to the entire dataset could inadvertently introduce biases or lead to over-fitting, undermining the model’s capacity to generalize to new and unseen data. Moreover, some of these methods may exact a computational and temporal cost, particularly when dealing with extensive datasets. Consequently, a prudent evaluation of these factors is crucial when determining the most suitable technique for a given project.

After conducting a thorough literature review, we have discovered that using a sampling technique on the entire raw data may result in data leakage and ultimately lead to inflated metric values (Tampu et al., 2022; Silva et al., 2022; Jagan Mohan et al., 2022; Jamuna Devi and Kavitha, 2023; Navarro et al., 2021). However, when appropriately used, our strategy enhances the outcome of severely imbalanced data compared to the initial and existing results (Xie et al., 2019). Also, it is imperative to note that comparing results to existing literature may only sometimes yield fruitful results because the parameters of the machine learning algorithm depend on the dataset’s unique characteristics, including the set of dependent and independent variables. As a result, changes in the dataset can affect the outcome(James et al., 2021; Rowe, 2019). Therefore, it is essential to consider how much improvement in prediction is happening compared to the raw data when making comparisons.

In summation, the implications of our study resonate deeply with the global burden of diabetes. We have built an end to end methodology for machine learning algorithms to handle the highly imbalance data. By facilitating early detection, enabling proactive interventions, and optimizing resource allocation, our holistic strategy seeks to transform the trajectory of diabetes prevention and management.

## Data Availability

https://www.cdc.gov/brfss/index.html

## Declaration of generative AI and AI-assisted technologies in the writing process

While preparing this work, the authors used https://chat.openai.com to improve language and readability. After using this tool, the authors reviewed and edited the content as needed and takes full responsibility for the publication’s content.

## References

R. S. Ahima. Connecting obesity, aging and diabetes. Nature medicine, 15(9):996–997, 2009.

R. Alejo, J. M. Sotoca, R. M. Valdovinos, and P. Toribio. Edited nearest neighbor rule for improving neural networks classifications. In Advances in Neural Networks-ISNN 2010: 7th International Symposium on Neural Networks, ISNN 2010, Shanghai, China, June 6-9, 2010, Proceedings, Part I 7, pages 303–310. Springer, 2010.

A. Anand, G. Pugalenthi, G. B. Fogel, and P. Suganthan. An approach for classification of highly imbalanced data using weighting and undersampling. Amino acids, 39:1385–1391, 2010.

D. Asiimwe, G. O. Mauti, and R. Kiconco. Prevalence and risk factors associated with type 2 diabetes in elderly patients aged 45-80 years at kanungu district. Journal of diabetes research, 2020:1–5, 2020.

A. D. Association. Diagnosis and classification of diabetes mellitus. Diabetes care, 33(Supplement 1): S62–S69, 2010.

A. D. Association. Economic costs of diabetes in the us in 2017. Diabetes care, 41(5):917–928, 2018.

A. D. Association. The cost of diabetes, Accessed June 22, 2023. American Diabetes Association Accessed https://diabetes.org/about-us/statistics/cost-diabetes.

A. Astrup and N. Finer. Redefining type 2 diabetes:’diabesity’or ‘obesity dependent diabetes mellitus’? Obesity reviews, 1(2):57–59, 2000.

G. Batista, R. Prati, and M.-C. Monard. A study of the behavior of several methods for balancing machine learning training data. SIGKDD Explorations, 6:20–29, 06 2004. doi: 10.1145/1007730.1007735.

M. Beyeler. Machine Learning for OpenCV. Packt Publishing Ltd, 2017.

J. J. Bigna and J. J. Noubiap. The rising burden of non-communicable diseases in sub-saharan africa. The Lancet Global Health, 7(10):e1295–e1296, 2019.

L. Breiman. Random forests. Machine learning, 45:5–32, 2001.

T. A. Buchanan, A. H. Xiang, et al. Gestational diabetes mellitus. The Journal of clinical investigation, 115(3):485–491, 2005.

A. Budreviciute, S. Damiati, D. K. Sabir, K. Onder, P. Schuller-Goetzburg, G. Plakys, A. Katileviciute, S. Khoja, and R. Kodzius. Management and prevention strategies for non-communicable diseases (ncds) and their risk factors. Frontiers in public health, page 788, 2020.

J. Burez and D. Van den Poel. Handling class imbalance in customer churn prediction. Expert Systems with Applications, 36(3):4626–4636, 2009.

C. J. Caspersen, G. D. Thomas, L. A. Boseman, G. L. Beckles, and A. L. Albright. Aging, diabetes, and the public health system in the united states. American journal of public health, 102(8): 1482–1497, 2012.

CDC. Behavioral risk factor surveillance system, Accessed March 22, 2023a. Center for Disease Control and Prevention Accessed at https://www.cdc.gov/brfss/index.html.

CDC. Diabetes basics, Accessed March 22, 2023b. Center for Disease Control and Prevention Accessed at https://www.cdc.gov/diabetes/basics/index.html.

CDC. Diabetes fast facts, Accessed March 22, 2023c. Center for Disease Control and Prevention Accessed at https://www.cdc.gov/diabetes/basics/quick-facts.html.

CDC. Diabetes and covid-19, Accessed March 22, 2023d. Center for Disease Control and Prevention Accessed at https://www.cdc.gov/diabetes/library/reports/reportcard/diabetes-and-covid19.html.

CDC. What is diabetes?, Accessed March 22, 2023e. Center for Disease Control Accessed https://www.cdc.gov/diabetes/basics/diabetes.html::text=Diabetes.

CDC. About prediabetes type 2 diabetes, Accessed March 22, 2023f. Center for Disease Control and Prevention Accessed at https://www.cdc.gov/diabetes/prevention/about-prediabetes.html.

CDC. Type 2 diabetes, Accessed March 22, 2023g. Accessed at https://www.cdc.gov/diabetes/basics/type2.html.

CDC. Global noncommunicable diseases fact sheet, Accessed March 24, 2023. Accessed at https://www.cdc.gov/globalhealth/healthprotection/resources/fact-sheets/global-ncd-fact-sheet.html::text=Noncommunicable.

S. Chatterjee, K. Khunti, and M. J. Davies. Type 2 diabetes. The lancet, 389(10085):2239–2251, 2017.

N. V. Chawla, K. W. Bowyer, L. O. Hall, and W. P. Kegelmeyer. Smote: synthetic minority over-sampling technique. Journal of artificial intelligence research, 16:321–357, 2002.

S. Chen, M. Kuhn, K. Prettner, and D. E. Bloom. The macroeconomic burden of noncommunicable diseases in the united states: Estimates and projections. PloS one, 13(11):e0206702, 2018.

M. M. Chowdhury, M. R. Islam, M. S. Hossain, N. Tabassum, and A. Peace. Incorporating the mutational landscape of sars-cov-2 variants and case-dependent vaccination rates into epidemic models. Infectious Disease Modelling, 7(2):75–82, 2022.

A. Cutler, D. R. Cutler, and J. R. Stevens. Random forests. Ensemble machine learning: Methods and applications, pages 157–175, 2012.

A. Dinh, S. Miertschin, A. Young, and S. D. Mohanty. A data-driven approach to predicting diabetes and cardiovascular disease with machine learning. BMC medical informatics and decision making, 19(1):1–15, 2019.

J. Divers, E. J. Mayer-Davis, J. M. Lawrence, S. Isom, D. Dabelea, L. Dolan, G. Imperatore, S. Marcovina, D. J. Pettitt, C. Pihoker, et al. Trends in incidence of type 1 and type 2 diabetes among youths—selected counties and indian reservations, united states, 2002–2015. Morbidity and Mortality Weekly Report, 69(6):161, 2020.

G. S. Eisenbarth. Type i diabetes mellitus. New England journal of medicine, 314(21):1360–1368, 1986.

A. Fernandez, S. Garcia, M. Galar, R. C. Prati, B. Krawczyk, and F. Herrera. Learning from imbalanced data sets, volume 10. Springer, 2018.

Y. Freund and R. E. Schapire. A decision-theoretic generalization of on-line learning and an application to boosting. Journal of Computer and System Sciences, 55(1):119–139, 1997. ISSN 0022-0000. 10.1006/jcss.1997.1504. URL https://www.sciencedirect.com/science/article/pii/S002200009791504X.

J. H. Friedman. Greedy function approximation: a gradient boosting machine. Annals of statistics, pages 1189–1232, 2001.

H. Frumkin and A. Haines. Global environmental change and noncommunicable disease risks. Annual review of public health, 40:261–282, 2019.

L. S. Geiss, D. B. Rolka, and M. M. Engelgau. Elevated blood pressure among us adults with diabetes, 1988–1994. American journal of preventive medicine, 22(1):42–48, 2002.

N. Gray, G. Picone, F. Sloan, and A. Yashkin. The relationship between bmi and onset of diabetes mellitus and its complications. Southern medical journal, 108(1):29, 2015.

M. H. Green. Taking” pandemic” seriously: Making the black death global. The Medieval Globe, 1 (1):27–61, 2015.

A. S. Group. Effects of intensive blood-pressure control in type 2 diabetes mellitus. New England Journal of Medicine, 362(17):1575–1585, 2010.

S. H. Habib and S. Saha. Burden of non-communicable disease: global overview. Diabetes and Metabolic Syndrome: Clinical Research and Reviews, 4(1):41–47, 2010.

T. Hastie, R. Tibshirani, J. H. Friedman, and J. H. Friedman. The elements of statistical learning: data mining, inference, and prediction, volume 2. Springer, 2009.

H. He and E. A. Garcia. Learning from imbalanced data. IEEE Transactions on knowledge and data engineering, 21(9):1263–1284, 2009.

F. Hill-Briggs, N. E. Adler, S. A. Berkowitz, M. H. Chin, T. L. Gary-Webb, A. Navas-Acien, P. L. Thornton, and D. Haire-Joshu. Social determinants of health and diabetes: a scientific review. Diabetes care, 44(1):258–279, 2021.

M. R. Islam, T. Oraby, A. McCombs, M. M. Chowdhury, M. Al-Mamun, M. G. Tyshenko, and C. Kadelka. Evaluation of the united states covid-19 vaccine allocation strategy. PloS one, 16 (11):e0259700, 2021.

N. Jagan Mohan, R. Murugan, and T. Goel. Deep learning for diabetic retinopathy detection: Challenges and opportunities. Next Generation Healthcare Informatics, pages 213–232, 2022.

C. A. James, K. M. Wheelock, and J. O. Woolliscroft. Machine learning: the next paradigm shift in medical education. Academic Medicine, 96(7):954–957, 2021.

P. Jamuna Devi and B. Kavitha. Data leakage and data wrangling in machine learning for medical treatment. Data Wrangling: Concepts, Applications and Tools, pages 91–107, 2023.

N. Japkowicz. Why question machine learning evaluation methods. In AAAI workshop on evaluation methods for machine learning, pages 6–11. Citeseer, 2006.

S. Kastora, M. Patel, B. Carter, M. Delibegovic, and P. K. Myint. Impact of diabetes on covid-19 mortality and hospital outcomes from a global perspective: An umbrella systematic review and meta-analysis. Endocrinology, Diabetes & Metabolism, 5(3):e00338, 2022.

A. Katsarou, S. Gudbjörnsdottir, A. Rawshani, D. Dabelea, E. Bonifacio, B. J. Anderson, L. M. Jacobsen, D. A. Schatz, and Å. Lernmark. Type 1 diabetes mellitus. Nature reviews Disease primers, 3(1):1–17, 2017.

H. Kaur, H. S. Pannu, and A. K. Malhi. A systematic review on imbalanced data challenges in machine learning: Applications and solutions. ACM Computing Surveys (CSUR), 52(4):1–36, 2019.

N. Kenworthy, M. Thomann, and R. Parker. From a global crisis to the ‘end of aids’: New epidemics of signification. Global Public Health, 13(8):960–971, 2018.

K. S. Leong and J. P. Wilding. Obesity and diabetes. Best Practice & Research Clinical Endocrinology & Metabolism, 13(2):221–237, 1999.

J. E. Morley. Diabetes and aging: epidemiologic overview. Clinics in geriatric medicine, 24(3): 395–405, 2008.

C. L. A. Navarro, J. A. Damen, T. Takada, S. W. Nijman, P. Dhiman, J. Ma, G. S. Collins, R. Bajpai, R. D. Riley, K. G. Moons, et al. Risk of bias in studies on prediction models developed using supervised machine learning techniques: systematic review. bmj, 375, 2021.

A. Rajpal, L. Rahimi, and F. Ismail-Beigi. Factors leading to high morbidity and mortality of covid-19 in patients with type 2 diabetes. Journal of diabetes, 12(12):895–908, 2020.

G. Robertson, E. D. Lehmann, W. Sandham, and D. Hamilton. Blood glucose prediction using artificial neural networks trained with the aida diabetes simulator: a proof-of-concept pilot study. Journal of Electrical and Computer Engineering, 2011:2–2, 2011.

M. Rowe. An introduction to machine learning for clinicians. Academic Medicine, 94(10):1433–1436, 2019.

P. Saeedi, I. Petersohn, P. Salpea, B. Malanda, S. Karuranga, N. Unwin, S. Colagiuri, L. Guariguata, A. A. Motala, K. Ogurtsova, et al. Global and regional diabetes prevalence estimates for 2019 and projections for 2030 and 2045: Results from the international diabetes federation diabetes atlas. Diabetes research and clinical practice, 157:107843, 2019.

V. Shriraam, S. Mahadevan, and P. Arumugam. Prevalence and risk factors of diabetes, hypertension and other non-communicable diseases in a tribal population in south india. Indian Journal of Endocrinology and Metabolism, 25(4):313, 2021.

G. F. Silva, T. P. Fagundes, B. C. Teixeira, and A. D. Chiavegatto Filho. Machine learning for hypertension prediction: a systematic review. Current Hypertension Reports, 24(11):523–533, 2022.

S. Supakul, H. Y. Park, B. N. Nguyen, and K. B. Giang. Prevalence differences in major non-communicable diseases in a low-middle income country: a comparative study between an urban and a rural district in vietnam. Journal of Global Health Science, 1(2), 2019.

I. E. Tampu, A. Eklund, and N. Haj-Hosseini. Inflation of test accuracy due to data leakage in deep learning-based classification of oct images. Scientific Data, 9(1):580, 2022.

I. Ul Hassan, R. H. Ali, Z. Ul Abideen, T. A. Khan, and R. Kouatly. Significance of machine learning for detection of malicious websites on an unbalanced dataset. Digital, 2(4):501–519, 2022.

Z. Ullah, F. Saleem, M. Jamjoom, B. Fakieh, F. Kateb, A. M. Ali, B. Shah, et al. Detecting high-risk factors and early diagnosis of diabetes using machine learning methods. Computational Intelligence and Neuroscience, 2022, 2022.

J. M. Van Seventer and N. S. Hochberg. Principles of infectious diseases: transmission, diagnosis, prevention, and control. International encyclopedia of public health, page 22, 2017.

WHO. Global health estimates: Life expectancy and leading causes of death and disability, Accessed March 22, 2023a. World Health Organization Accessed at https://www.who.int/data/gho/data/themes/mortality-and-global-health-estimates.

WHO. Noncommunicable diseases, Accessed March 22, 2023b. World Health Organization Accessed at https://www.who.int/news-room/fact-sheets/detail/noncommunicable-diseases.

WHO. Noncommunicable diseases, Accessed March 22, 2023c. World Health Organization Accessed at https://www.who.int/news-room/fact-sheets/detail/noncommunicable-diseases.

R. Williams, S. Karuranga, B. Malanda, P. Saeedi, A. Basit, S. Besançon, C. Bommer, A. Esteghamati, K. Ogurtsova, P. Zhang, et al. Global and regional estimates and projections of diabetes-related health expenditure: Results from the international diabetes federation diabetes atlas. Diabetes research and clinical practice, 162:108072, 2020.

D. L. Wilson. Asymptotic properties of nearest neighbor rules using edited data. IEEE Transactions on Systems, Man, and Cybernetics, (3):408–421, 1972.

Y. Wu, Y. Ke, Z. Chen, S. Liang, H. Zhao, and H. Hong. Application of alternating decision tree with adaboost and bagging ensembles for landslide susceptibility mapping. Catena, 187:104396, 04 2020. doi: 10.1016/j.catena.2019.104396.

Z. Xie, O. Nikolayeva, J. Luo, and D. Li. Peer reviewed: building risk prediction models for type 2 diabetes using machine learning techniques. Preventing chronic disease, 16, 2019.

A. Zumla, A. N. Alagaili, M. Cotten, and E. I. Azhar. Infectious diseases epidemic threats and mass gatherings: refocusing global attention on the continuing spread of the middle east respiratory syndrome coronavirus (mers-cov). BMC medicine, 14(1):1–4, 2016.

